# “Mental health status of the general population, healthcare professionals, and university students during 2019 coronavirus disease outbreak in Jordan: a cross-sectional study”

**DOI:** 10.1101/2020.04.09.20056374

**Authors:** Abdallah Y Naser, Eman Zmaily Dahmash, Rabaa Al-Rousan, Hassan Alwafi, Hamzeh Mohammad Alrawashdeh, Imene Ghoul, Anwer Abidine, Mohammed A. Bokhary, Hadeel T. AL-Hadithi, Dalia Ali, Rasha Abuthawabeh, Ghada Mohammad Abdelwahab, Yosra J Alhartani, Haneen Al Muhaisen, Ayah Dagash

## Abstract

**Background:** The emergence of COVID-19 global pandemic coupled with high transmission rate and mortality has created an unprecedented state of emergency worldwide. This global situation may have a negative impact on the psychological well-being of individuals which in turn impacts individuals’ performance.

**Methods:** A cross-sectional study using an online survey was conducted in Jordan between 22nd and 28th of March 2020 to explore the mental health status (depression and anxiety) of the general population, healthcare professionals, and university students during the COVID-19 outbreak. The Patient Health Questionnaire (PHQ-9) and Generalized Anxiety Disorder-7 (GAD-7) were used to assess depression and anxiety among the study participants. Logistic regression analysis was used to identify predictors of depression and anxiety.

**Results:** The prevalence of depression and anxiety among the entire study participants was 23.8% and 13.1%, respectively. Anxiety was most prevalent across university students 21.5%, followed by healthcare professionals 11.3%, and general population 8.8%. Females among healthcare professionals and university students, divorced healthcare professionals, pulmonologists, and university students with history of chronic disease were at higher risk of developing depression. Females, divorced participants among the general population, and university students with history of chronic disease and those with high income (≥1500 JD) were at higher risk of developing anxiety.

**Conclusions:** During outbreaks, individuals are put under extreme stressful condition resulting in higher risk of developing anxiety and depression particularly for students and healthcare professionals. Policymakers and mental healthcare providers are advised to provide further mental support to these vulnerable groups during this pandemic.

## 1. INTRODUCTION

In December 2019, an infectious disease with unknown aetiology characterised with acute pneumonia has been recognized in Wuhan, China, named 2019 coronavirus disease (COVID-19) (Wang et al., 2020). The causative microorganism has been identified as a new RNA virus from the beta-coronavirus family, named as severe acute respiratory syndrome coronavirus 2 (SARS-CoV-2) (Guan et al., 2020). The respiratory illness caused by the COVID-19 is highly contagious, therefore, the World Health Organization (WHO) has categorised it as a pandemic infection (WHO, 2020).

COVID-19 is mainly transmitted through respiratory droplets and close contact, making a huge number of the population at risk of getting infected. The widespread of COVID-19 mainly in Wuhan, China and worldwide has attracted attention all over the world. As of April 06, 2020, a total of 1,254,969 persons were diagnosed with COVID-19 and 68,825 died of this life-threating infectious disease (Channel news Asia, 2020). In Jordan, a total of 345 cases were confirmed and 110 patients recovered with five deaths reported (Channel news Asia, 2020; World Stats, 2020).

Due to duty calls, healthcare professionals (HCP) are more likely to come in contact with COVID-19 carriers putting them at a greater risk of contracting the infection and spreading it to others, including their loved ones. A recent study in China reported that a total of 2,055 HCPs were diagnosed with COVID-19 (National Health Commission, 2020). In response to COVID-19 pandemic, Jordan government took extremely proactive measures to prevent the spread of the virus where the state of emergency was declared on March 20, 2020. The country witnessed a complete lockdown, banning people from leaving their homes except for a few specific reasons. Individual violating these orders were subject to three years in prison, a $4,200 fine, or both. The government also implemented an extensive public health awareness campaign utilizing all available media channels (The Jordan Times, 2020). The exponential increase in the number of cases and deaths in better-equipped countries, the spread of COVID-19 misinformation, the lack of medical treatment, and the shortage of properly equipped units to care for patients all could contribute to provoking public fear, anxiety and/or depression, which is usually neglected during crisis and pandemic management (Cao et al., 2020; Chen et al., 2020; Downes, 2015; Huang & Zhao, 2020; Zhai & Du, 2020).

Psychological wellbeing has an important impact on individuals’ performance. This impact is well documented among different populations including HCPs (Ramirez, Graham, Richards, Gregory, & Cull, 1996; Schafheutle, Seston, & Hassell, 2011; Wallace, Lemaire, & Ghali, 2009), general population (GP) (Burt, Zembar, & Niederehe, 1995), and university students (US) (Andrews & Wilding, 2004; Bruffaerts et al., 2018; Campos, Oliveira, Mello, & Dantas, 2017; O’Malley, Voight, Renshaw, & Eklund, 2015). Therefore, the study of the psychological wellbeing of these populations during COVID-19 pandemic is of paramount importance. This study aims to explore the prevalence of depression and anxiety among the GP, HCPs, and USs during COVID-19 outbreak, and identify key population(s) who might need psychological intervention.

## 2. METHODS

### 2.1. Study design and study population

A cross-sectional study by means of online survey was conducted in Jordan between 22^nd^ and 28^th^ of March 2020 to explore the mental health status (depression and anxiety) of the GP, HCPs, and USs during the outbreak of COVID-19. The data collection period was restricted to one week as longer period may influence the mental-health status of the study populations.

#### 2.1.1 Sampling strategy

A convenience sample of eligible participants was invited to participate in the study. The GP, HCPs, and USs were invited to participate in this study through social media (Facebook and WhatsApp). Each study population was invited using a specific survey-link as each population has specific demographic questions. All participants voluntarily participated in the study and were thus considered exempt from written informed consent. Study aim and objectives were clearly explained at the beginning of the survey.

The inclusion criteria were: a) participants aged 18 years and above and living currently in Jordan; and b) participants who had no apparent cognitive deficit. Participants were excluded if they were: a) below 18 years of age; b) unable to understand Arabic language; and c) unable to participate due to physical or emotional distress.

#### 2.1.2. Depression and anxiety assessment scales

Previously validated assessment scales, the Patient Health Questionnaire (PHQ)-9 and Generalized Anxiety Disorder 7-item (GAD-7) were used to assess depression and anxiety among the study participants. These screening instruments were frequently used and validated as brief screening tools among various populations for depression and anxiety (Levis, Benedetti, Thombs, & Collaboration., 2019; Löwe et al., 2008; Martin, Rief, Klaiberg, & Braehler, 2006; Maurer, Raymond, & Davis, 2018; S. Yoon et al., 2014). In addition, the following information were collected: participants’ demographics (age, gender, income, and marital status). Furthermore, all participants were asked whether they were worried about being infected with COVID-19 or transmitting it to family members (yes/no question). The GP and USs were asked whether they had underlying chronic conditions (yes/no question). The GP were also asked about their education level and employment status. HCPs were asked about their specialities and exposure to COVID-19 patients and/or providing medical care for COVID-19 patients during this pandemic. USs were asked about their field of study and year level.

The PHQ-9 scale is a 9-questions instrument given to participants to screen for the presence and severity of depression (Hartung et al., 2017; Hinz et al., 2016). The GAD-7 instrument was used to screen for anxiety (Esser et al., 2018). The PHQ-9 and the GAD-7 instruments asked the participants about the degree of applicability of each item (question), using a 4-point Likert scale. Participants’ response ranged from 0 to 3, where 0 means “Not at all” and 3 means “Nearly every day”. The PHQ-9 instrument includes 9 items. Items are scored from 0 to 3 generating a total score ranging from 0 to 27. A total score of 0–4 indicates minimal depression, 5–9 mild depression, 10–14 moderate depression, 15-19 moderately severe depression, and 20–27 severe depression (Schwenk et al., 2011). The GAD-7 instrument includes 7 items. Items are scored from 0 to 3 generating a total score ranging from 0 to 21. A total score of 5–9 indicates mild anxiety, 10–14 moderate anxiety, and 15–21 severe anxiety (Spitzer, Kroenke, Williams, & Löwe, 2006).

#### 2.1.3. Estimate of prevalence and classification of depression and anxiety

Prevalence rates of depression and anxiety were determined using a cut-off point as recommended by the authors of the PHQ-9 and GAD-7 scale. In this study, depression was defined as a total score of (≥ 15) in the PHQ-9 instrument indicating a case with moderately severe or severe depression. Anxiety was defined using the GAD-7 instrument with a total score of (≥ 15) indicating a case with severe anxiety. The higher the score the more severe the case identified by any scale.

The prevalence rate of depression was estimated by dividing the number of participants who exceeded the borderline score (≥ 15) by the total number of the participants in the same population. Prevalence rate of anxiety was calculated using the same procedure.

#### 2.1.4. Sample size

The target sample size was estimated based on the WHO recommendations for the minimal sample size needed for a prevalence study (Lwanga & Lemeshow, 1991). Using a confidence interval of 95%, a standard deviation of 0.5, a margin of error of 5%, the required sample size was 385 participants from each study population.

#### 2.1.5. Ethical approval

This study was approved by the Research Ethics Committee at Faculty of Pharmacy in Isra University, Amman, Jordan. As participation in the study was voluntary, the Research Ethics Committee approved consent waiver.

### 2.2. Statistical analysis

Descriptive statistics were used to describe participants’ demographic characteristics. Continuous data were reported as mean ± SD for normally distributed variables and median (interquartile range (IQR)) for non-normally distributed variables. Categorical data were reported as percentages (frequencies). The Mann-Whitney U test/Kruskal-Wallis test was used to compare the median scores between different demographic groups. Logistic regression was used to estimate odds ratios (ORs) with 95% confidence intervals (CIs) for anxiety or depression. Logistic regression models were carried out using anxiety or depression scores above the cut-off points highlighted above. A two-sided p<0.05 was considered as statistically significant. The statistical analyses were carried out using SPSS (version 25).

## 3. Results

### 3.1. Participants characteristics

A total of 4,126 individuals participated in the study (GP=1,798, HCPs=1,163, and USs=1,165). **Table 1** details the baseline characteristics of the participants in the three populations. The majority of participants (n=2,436, 59.0%) were females, aged between 18–29 years (n=2,287, 55.4%), single (n=2,225, 53.9%), and with an income of 500 JD or below (n=2,152, 52.2%). Around half of the GP were employed (n=927, 51.6%), with the majority (n=1,190, 66.2%) completed their bachelor’s degree. The largest proportion of the participating HCPs were physicians (n=560, 48.2%). More than half of the participating USs (n=621, 53.3%) were studying non-medical sciences.

**TABLE 1.**
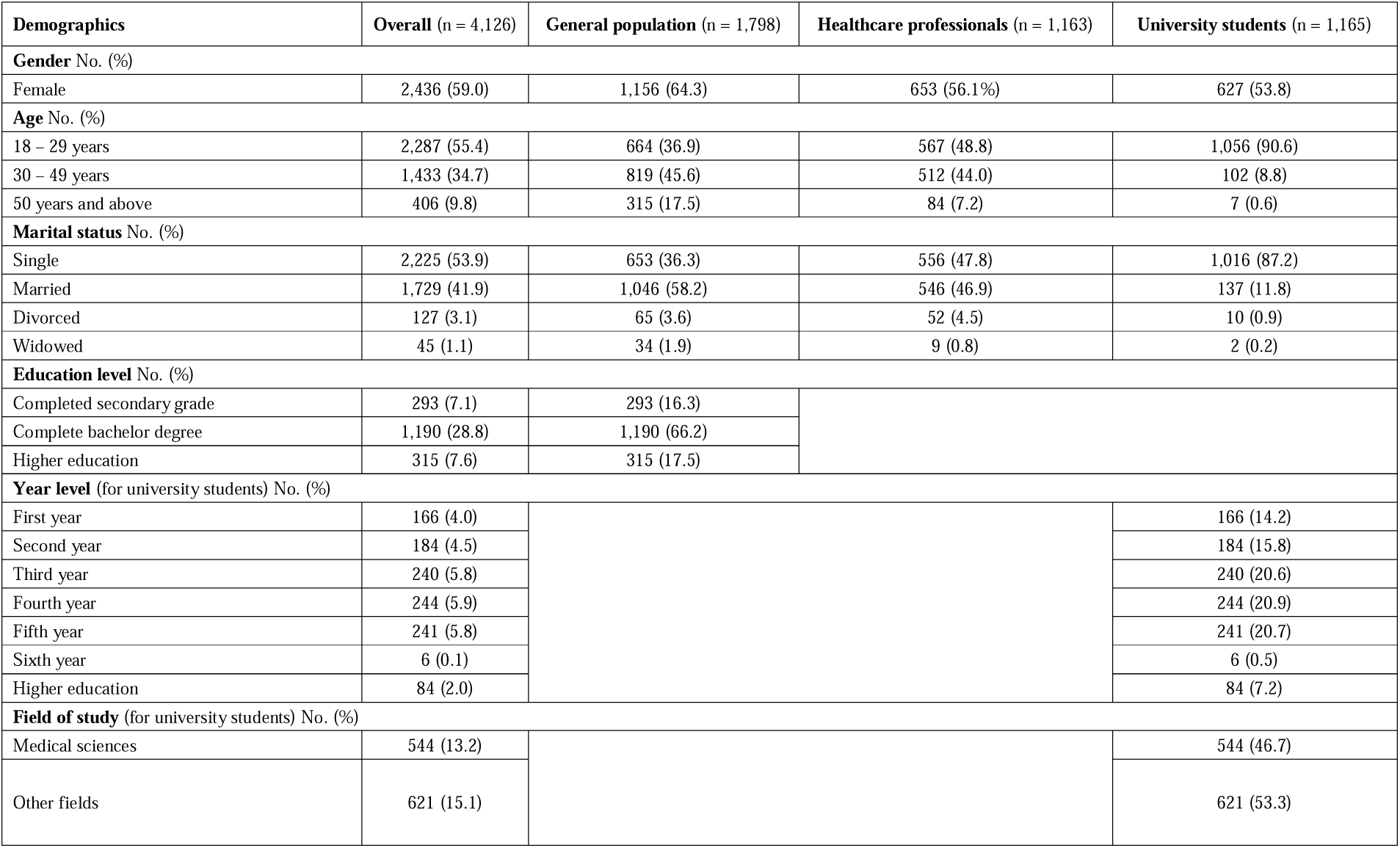
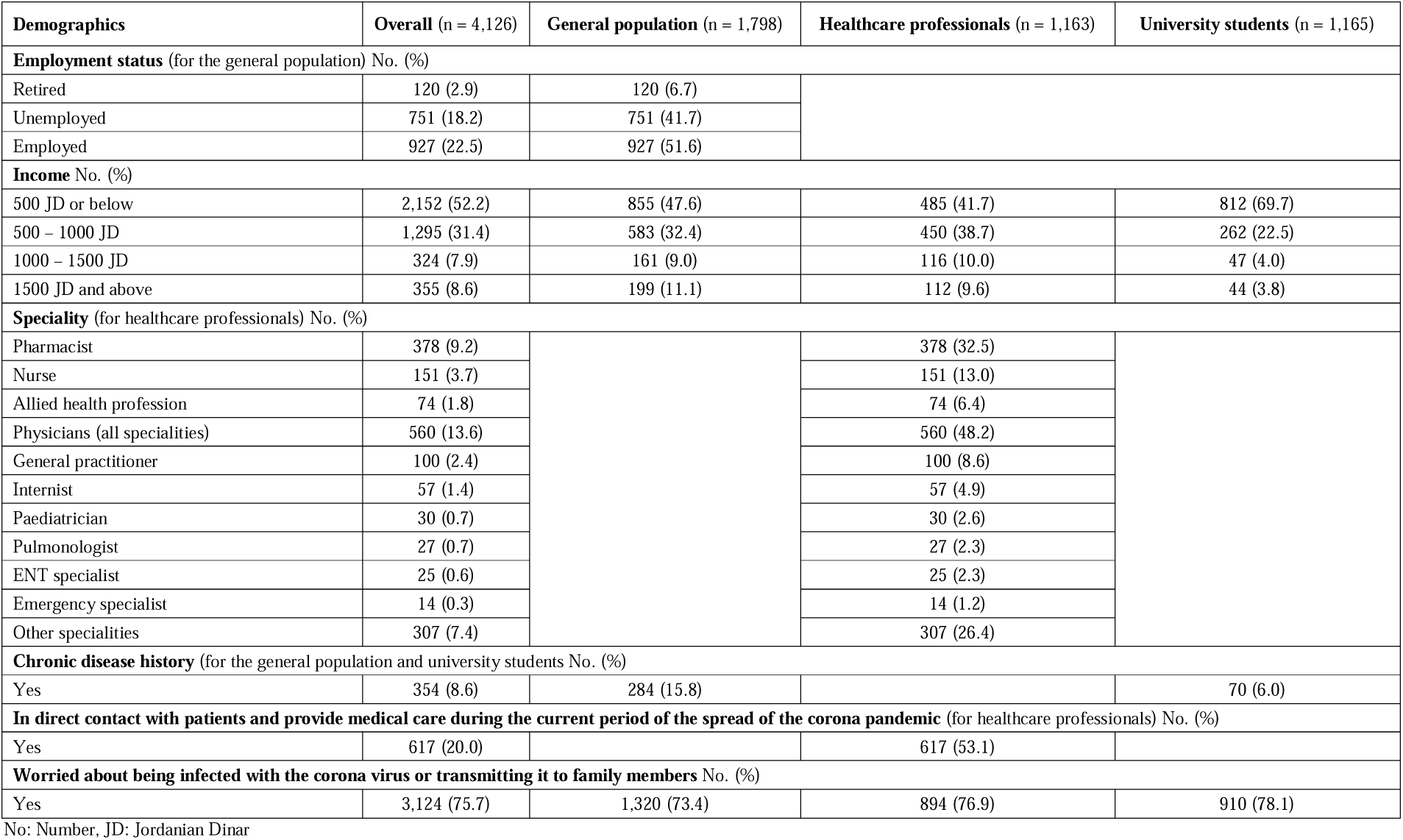
Participants characteristics from each population

| Demographics | Overall (n = 4,126) | General population (n = 1,798) | Healthcare professionals (n = 1,163) | University students (n = 1,165) |
| --- | --- | --- | --- | --- |
| Gender No. (%) |  |  |  |  |
| Female | 2,436 (59.0) | 1,156 (64.3) | 653 (56.1%) | 627 (53.8) |
| Age No. (%) |  |  |  |  |
| 18 – 29 years | 2,287 (55.4) | 664 (36.9) | 567 (48.8) | 1,056 (90.6) |
| 30 – 49 years | 1,433 (34.7) | 819 (45.6) | 512 (44.0) | 102 (8.8) |
| 50 years and above | 406 (9.8) | 315 (17.5) | 84 (7.2) | 7 (0.6) |
| Marital status No. (%) |  |  |  |  |
| Single | 2,225 (53.9) | 653 (36.3) | 556 (47.8) | 1,016 (87.2) |
| Married | 1,729 (41.9) | 1,046 (58.2) | 546 (46.9) | 137 (11.8) |
| Divorced | 127 (3.1) | 65 (3.6) | 52 (4.5) | 10 (0.9) |
| Widowed | 45 (1.1) | 34 (1.9) | 9 (0.8) | 2 (0.2) |
| Education level No. (%) |  |  |  |  |
| Completed secondary grade | 293 (7.1) | 293 (16.3) |  |  |
| Complete bachelor degree | 1,190 (28.8) | 1,190 (66.2) |  |  |
| Higher education | 315 (7.6) | 315 (17.5) |  |  |
| Year level (for university students) No. (%) |  |  |  |  |
| First year | 166 (4.0) |  |  | 166 (14.2) |
| Second year | 184 (4.5) |  |  | 184 (15.8) |
| Third year | 240 (5.8) |  |  | 240 (20.6) |
| Fourth year | 244 (5.9) |  |  | 244 (20.9) |
| Fifth year | 241 (5.8) |  |  | 241 (20.7) |
| Sixth year | 6 (0.1) |  |  | 6 (0.5) |
| Higher education | 84 (2.0) |  |  | 84 (7.2) |
| Field of study (for university students) No. (%) |  |  |  |  |
| Medical sciences | 544 (13.2) |  |  | 544 (46.7) |
| Other fields | 621 (15.1) |  |  | 621 (53.3) |
| Employment status (for the general population) No. (%) |  |  |  |  |
| Retired | 120 (2.9) | 120 (6.7) |  |  |
| Unemployed | 751 (18.2) | 751 (41.7) |  |  |
| Employed | 927 (22.5) | 927 (51.6) |  |  |
| Income No. (%) |  |  |  |  |
| 500 JD or below | 2,152 (52.2) | 855 (47.6) | 485 (41.7) | 812 (69.7) |
| 500 – 1000 JD | 1,295 (31.4) | 583 (32.4) | 450 (38.7) | 262 (22.5) |
| 1000 – 1500 JD | 324 (7.9) | 161 (9.0) | 116 (10.0) | 47 (4.0) |
| 1500 JD and above | 355 (8.6) | 199 (11.1) | 112 (9.6) | 44 (3.8) |
| Speciality (for healthcare professionals) No. (%) |  |  |  |  |
| Pharmacist | 378 (9.2) |  | 378 (32.5) |  |
| Nurse | 151 (3.7) |  | 151 (13.0) |  |
| Allied health profession | 74 (1.8) |  | 74 (6.4) |  |
| Physicians (all specialities) | 560 (13.6) |  | 560 (48.2) |  |
| General practitioner | 100 (2.4) |  | 100 (8.6) |  |
| Internist | 57 (1.4) |  | 57 (4.9) |  |
| Paediatrician | 30 (0.7) |  | 30 (2.6) |  |
| Pulmonologist | 27 (0.7) |  | 27 (2.3) |  |
| ENT specialist | 25 (0.6) |  | 25 (2.3) |  |
| Emergency specialist | 14 (0.3) |  | 14 (1.2) |  |
| Other specialities | 307 (7.4) |  | 307 (26.4) |  |
| Chronic disease history (for the general population and university students No. (%)) |  |  |  |  |
| Yes | 354 (8.6) | 284 (15.8) |  | 70 (6.0) |
| In direct contact with patients and provide medical care during the current period of the spread of the corona pandemic (for healthcare professionals) No. (%) |  |  |  |  |
| Yes | 617 (20.0) |  | 617 (53.1) |  |
| Worried about being infected with the corona virus or transmitting it to family members No. (%) |  |  |  |  |
| Yes | 3,124 (75.7) | 1,320 (73.4) | 894 (76.9) | 910 (78.1) |

No: Number, JD: Jordanian Dinar

Around 15.8% (n= 284) and 6.0% (n=70) of the GP and the USs respectively reported that they have a history of chronic disease. The vast majority (n=3,124, 75.7%) of the participants from all populations reported that they were concerned about contracting COVID-19 or transmitting it to family members. When participants were asked if they have identified any problems over the past two weeks, to what extent have these problems prevented them from doing their work, looking after their household affairs or dealing with people, 59.0% (n=2,436) reported that they faced difficulties.

### 3.2. Prevalence of mental health problems

The prevalence of depression (participants with a total score of 15 and above; cases with moderately severe and severe depression) among the entire study participants was 23.8% (n=980). Depression was most prevalent across USs (n=449, 38.6%), followed by HCPs (n=247, 21.2%) and GP (n=284, 15.8%). The proportions of minimal, mild, moderate, moderately severe, and severe depression were 24.3%, 31.8%, 20.2%, 13.4%, and 10.3% respectively. The prevalence of anxiety (participants with a total score of 15 and above; cases with severe anxiety) from the entire study participants was 13.1% (n=539). Similarly, anxiety was most prevalent across USs (n=250, 21.5%), followed by HCPs (n=131, 11.3%) and GP (n=158, 8.8%). The proportions of mild, moderate, and severe anxiety were 35.6%, 19.0%, and 13.1% respectively. **Table 2** details the prevalence of depression and anxiety among participants stratified by severity.

**TABLE 2.** Prevalence of depression and anxiety among the participants stratified by severity

|  | **Overall** (n = 4,126) | **General population**<br>(n = 1,798) | **Healthcare professionals**<br>(n = 1,163) | **University students**<br>(n = 1,165) |
| --- | --- | --- | --- | --- |
|  | **Depression diagnose** |  |  |  |
| **Minimal depression** | 1,003 (24.3) | 599 (33.3) | 256 (22.0) | 148 (12.7) |
| **Mild depression** | 1,311 (31.8) | 622 (34.6) | 387 (33.3) | 302 (25.9) |
| **Moderate depression** | 832 (20.2) | 293 (16.3) | 273 (23.5) | 266 (22.8) |
| **Moderately severe depression** | 554 (13.4) | 166 (9.2) | 152 (13.1) | 236 (20.3) |
| **Severe depression** | 426 (10.3) | 118 (6.6) | 95 (8.2) | 213 (18.3) |
|  | **Anxiety diagnose** |  |  |  |
| **Mild anxiety** | 1,469 (35.6) | 633 (35.2) | 442 (38.0) | 394 (33.8) |
| **Moderate anxiety** | 786 (19.0) | 252 (14.0) | 250 (21.5) | 284 (24.4) |
| **Severe anxiety** | 539 (13.1) | 158 (8.8) | 131 (11.3) | 250 (21.5) |

### 3.3. Participant demographics and mental health problems

**Table 3** presents participant demographics data and their median depression and anxiety scores. Depression median score significantly differed across participants from different demographical characteristics (p<0.05), except for the year level (for USs population). Anxiety median score significantly differed across participants from different demographical characteristics (p<0.01), except for educational level (for the GP), year level and field of study (for USs population).

**TABLE 3.**
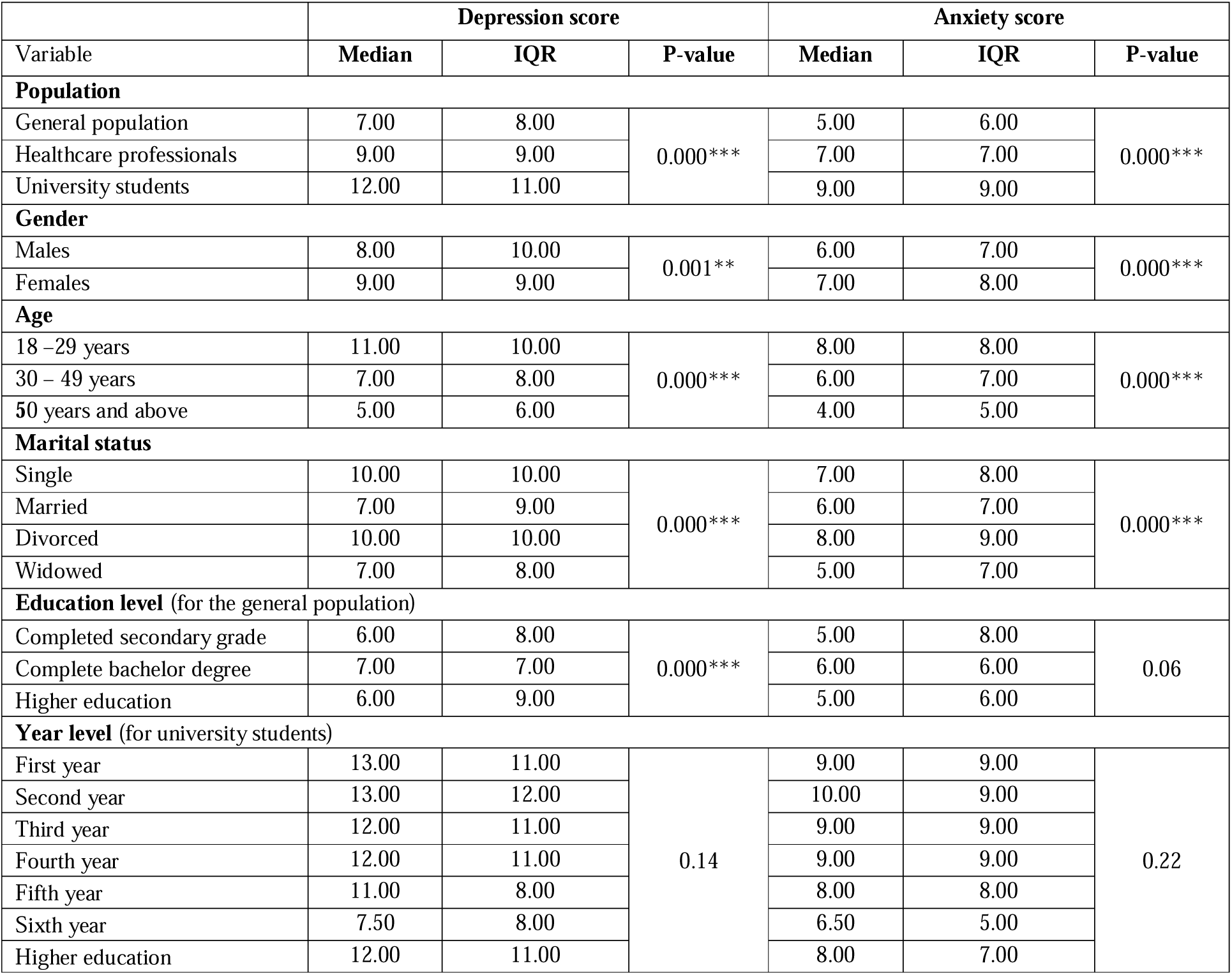
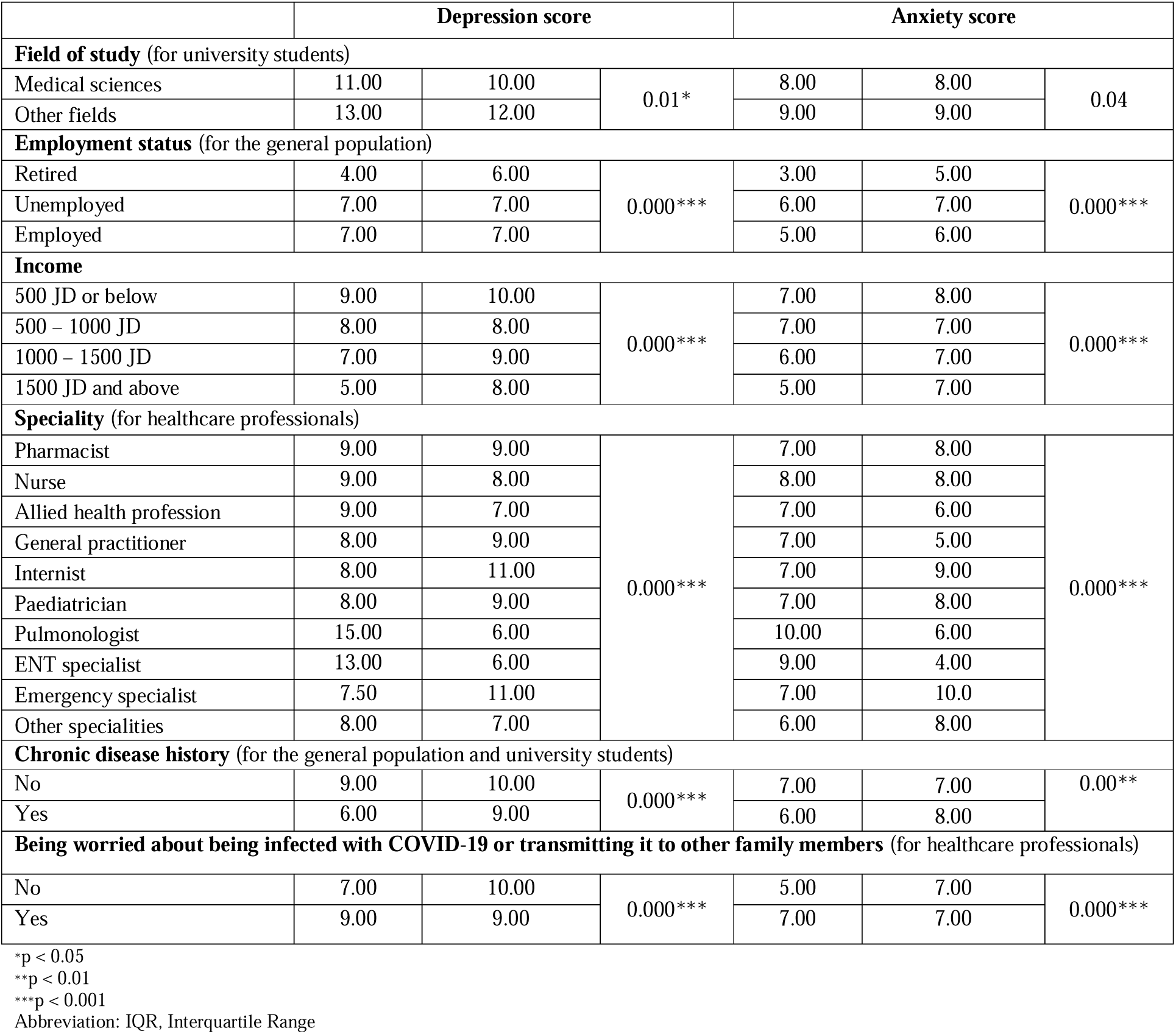
Depression and anxiety median score stratified by participants’ characteristics

University students, females, younger population (18–29 years), single and divorced, participants who completed their bachelor degree (from the GP), with lower-income category (500 JD and below), pulmonologist and ENT specialists (for HCPs), participants with no history of chronic diseases (for the GP and USs) tend to have higher depression and anxiety median scores compared to others.

The logistic regression analysis identified the following group to be at a higher risk of depression: a) females among HCPs and USs, b) divorced HCPs, c) pulmonologist, and d) USs with chronic disease history. On the other hand, the following groups were at a lower risk of depression: a) elderly, married, and high income (1500 JD and above) participants among the GP and HCPs, b) students at their fifth year of study, and c) retried participants from the GP. In addition, logistic regression analysis showed that the following groups were at a higher risk of anxiety: a) females across the three study populations, b) divorced participants among the GP, and c) USs with chronic disease history and who are with high income (1500 JD and above). On the other hand, the following groups were at a lower risk of anxiety: a) elderly and married participants among the GP, b) HCPs with high income (1500 JD and above) **Table 4**.

**TABLE 4.**
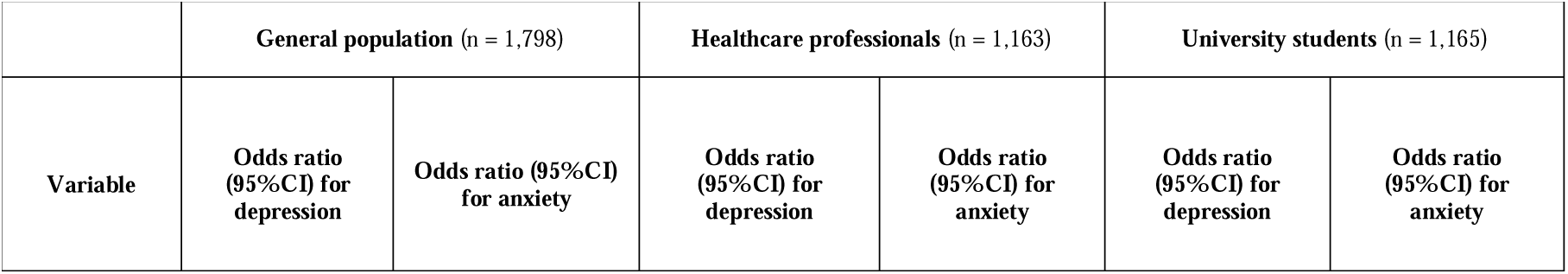
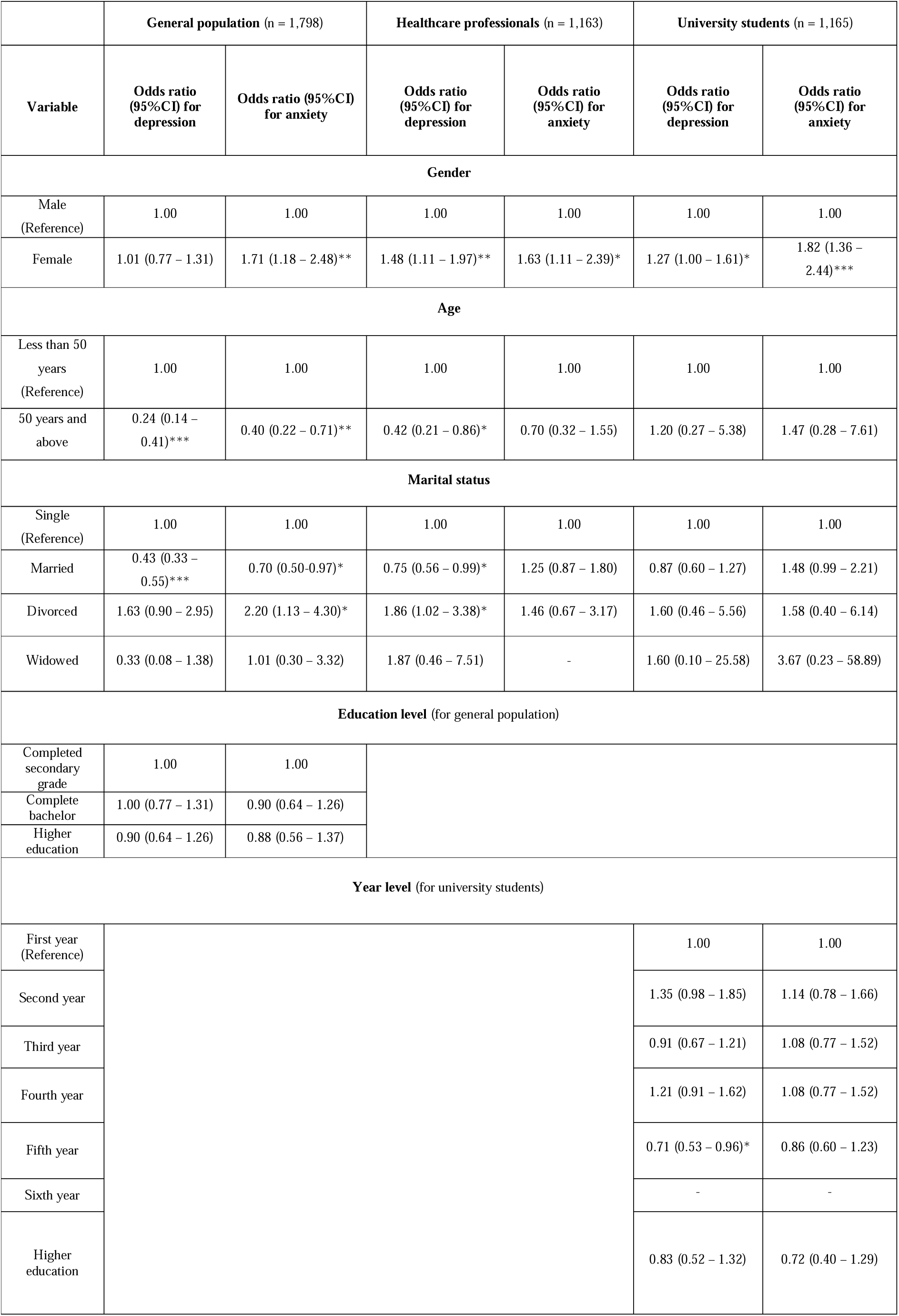
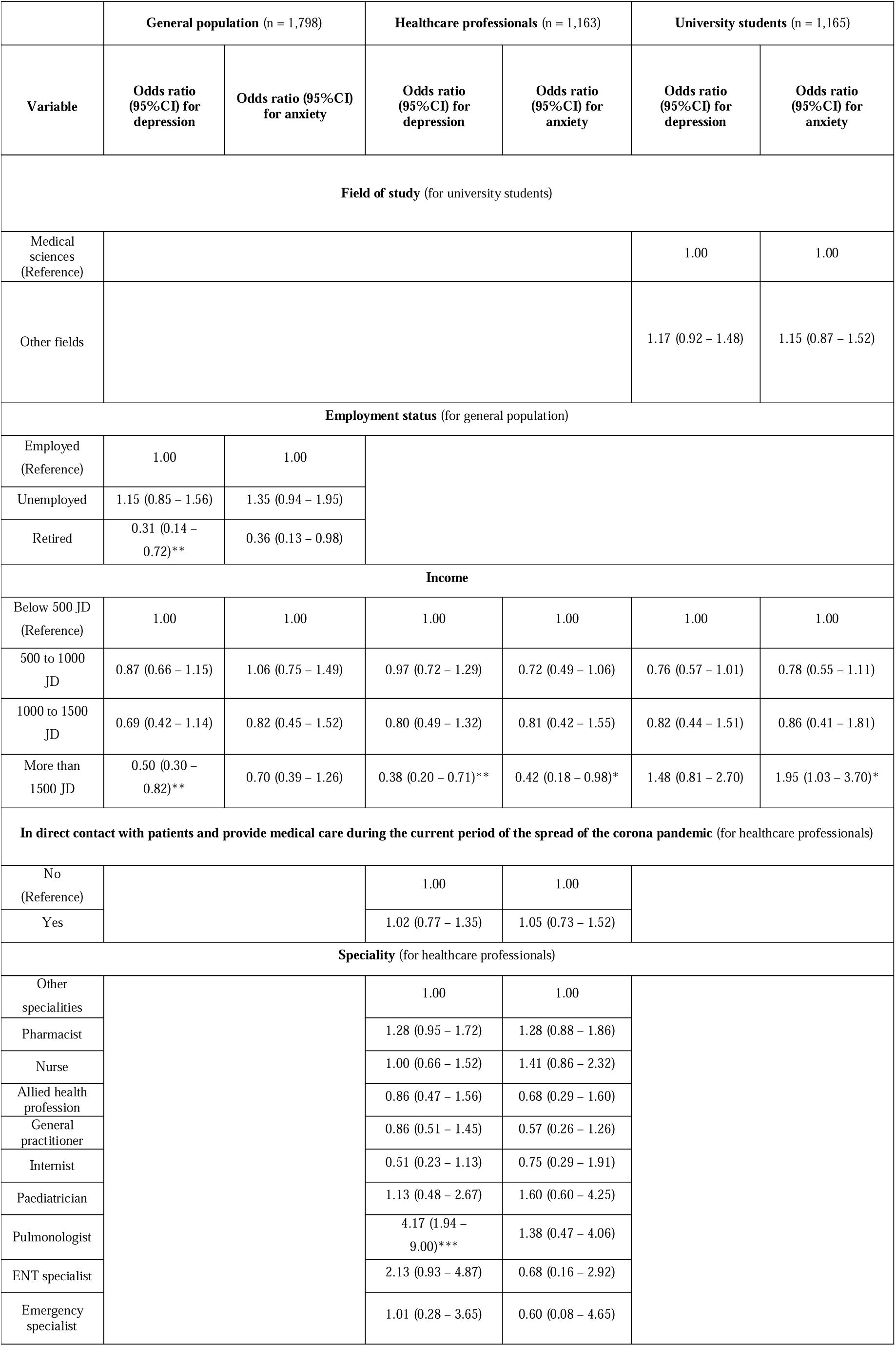
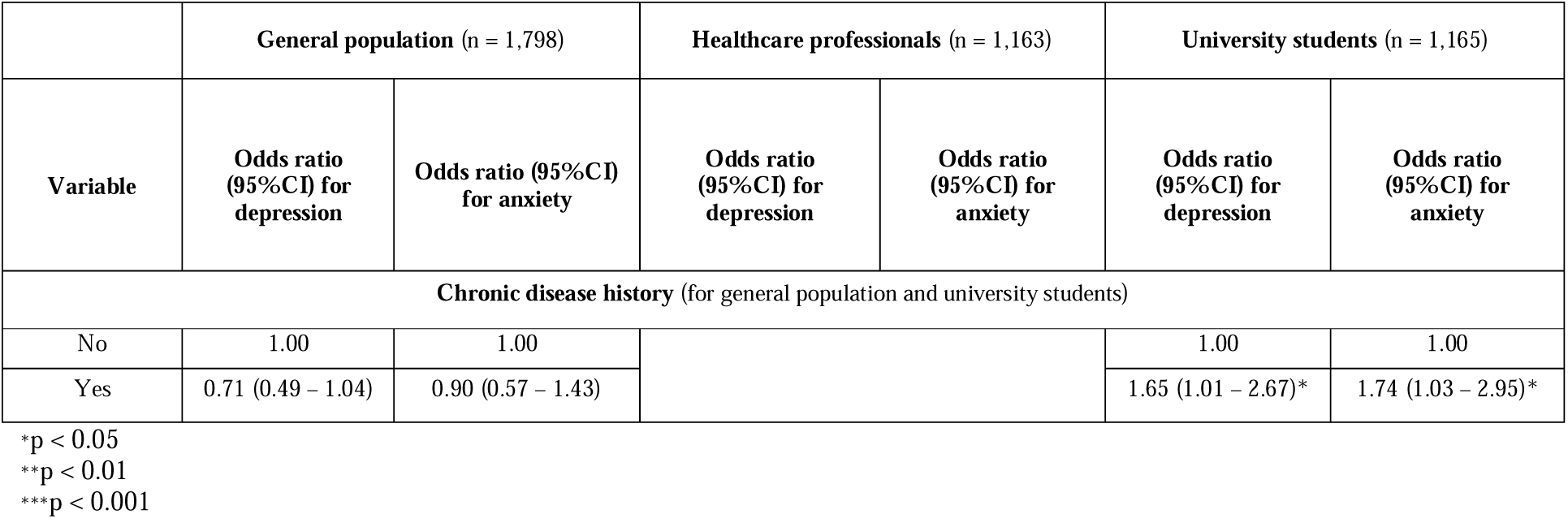
Logistic regression analysis

## 4. Discussion

Emerging COVID-19 is a recent pandemic the has exhausted the world’s resources including the lives of many. Therefore, studies that investigated the impact of this novel pandemic on mental health are limited. A recent study in China investigated the effect of COVID-19 on public psychological status, and reported that females were three-times at a higher risk of developing anxiety. On the other hand, higher education level was associated with a lower risk of developing depression by 0.39 time, these results are in-line with our study findings (Wang, Di, Ye, & Wei, 2020). Another study with 52,730 participants revealed that 35.0% of respondents experienced psychological distress during the epidemic with female respondents showing a higher risk of psychological distress when compared to males. Similarly, our results showed that females demonstrated a higher risk of anxiety and depression (p<0.05) which is in-line with reported data during epidemics, where women, particularly those working in healthcare, were prone to developing depression and anxiety (Lai et al., 2020; Li L, 2003). These gender differences in psychological distress including depression and anxiety have been described previously. Several studies have demonstrated that female gender is a risk factor for developing mental illness where women have been reported to have 1.6 greater incidence of depression compared to men. This could be attributed to increased frequency of hormonal fluctuation in women compared to men (Albert, 2015; Bartels, Cacioppo, van Beijsterveldt, & Boomsma, 2013).

In this study, a significant proportion of all participants (75.7%) emphasized their concerns about contracting COVID-19 or transmitting it to family members. Interestingly, although elderly are at higher risk of developing COVID-19 complications, our findings demonstrated that elderly participants (≥50 years) within the GP group had a significant lower risk of developing depression (OR for >50 years = 0.24, 95% CI 0.14 – 0.41, p<0.000) and anxiety (OR for >50 years = 0.40, 95% CI 0.22 – 0.7, p<0.01) compared to younger population. Possible explanation for these findings include lack of knowledge about possible complications, faith, or submission to mortality.

On the other hand, mental problems were most prevalent among USs showing 38.5%, 21.5% for depression and anxiety, respectively, followed by HCPs (21.2%, 11.3% respectively). The percentage of students suffering from depression and anxiety is alarming. This high prevalence rate could be primarily attributed to the mandatory switch to distance education despite the limited resources and skillsets. Therefore, students had major concerns regarding the impact of this emergency situation on their education and overall school performance. Interestingly, when students’ category was stratified according to year level, there was a slightly higher depression and anxiety score among first- and second-year students, however, it was statistically non-significant (p>0.05). This result tallied with the findings that a statistically significant higher prevalence rate of depression and anxiety was among young adults (18–29 years, p<0.05). Several studies have investigated the association between healthcare job-related stress with depression and anxiety (Gao et al., 2012; Mann & Cowburn, 2005; Smolders et al., 2009; S. L. Yoon & Kim, 2013) and such association appears more pronounced during a highly transmissible global pandemic such as COVID-19 (Lai et al., 2020). Our findings demonstrated that HCPs have a higher risk of developing unfavourable mental health outcomes particularly depression and anxiety. As we anticipated, pulmonologists and ENT physicians scored the highest in comparison to other HCPs. Potential factors that has contributed to these findings include pulmonologists and ENT physicians being in the front-line in treating COVID-19 patients, physician burnout, isolation form family, and feeling helpless due to the nature of this disease. Similar findings are reported by Wong and colleagues (2007) during SARS outbreak where ER physicians were found to have feeling of vulnerability, loss of control, fear of being infected or spreading the virus to family members and others (Wong et al., 2007). The unclear dynamics of COVID-19 transmission coupled with the high morbidity and mortality may exaggerate the perception of individual danger and provoke depression and anxiety (W. Wang, Tang, & Wei, 2020).

Although ENT physician score for depression and anxiety were statistically non-significant, which could be attributed to small sample size, we strongly believe that they are clinically important and should not be ignored. (p>0.05). Furthermore, foreseeable shortages of medical supplies, personal protective equipment (PPE) and the increasing number of both suspected and confirmed cases of COVID-19 contribute to the pressures and anxiety for HCPs (Chan-Yeung, 2004).

In comparison to USs and HCPs, the GP exhibited a lower prevalence rate of depression and anxiety. Possible contributor to these findings include the faith that Jordanians have in government. A study conducted by Analyseize Research demonstrated that 95.0% of Jordanians have full faith in government emergency measures to combat COVID-19 outbreak (Analyseize Research, 2020). Of note, married individuals were found to be at a lower risk of developing depression; GPs (OR= 0.43, 95% CI 0.33-0.55) and HCPs (OR= 0.75, 95% CI 0.56-0.99). Previous research reported that married couples have less tendency to develop depression and anxiety compared with divorced individuals which could be attributed to the impact of marriage on wellbeing and partner support (Kalmijn & Monden, 2006). Furthermore, our results showed that retired individuals among the GP tend to have a lower risk of developing depression (OR= 0.31, 95% CI 0.14-0.72. p<0.01) when compared to employed or unemployed individuals, which is consistent with previous studies. On one research conducted by Buffel and co-worker, they reported that retired individuals tend to have a lower score of depression during crisis when compared to employed and unemployed persons (Buffel, Van de Velde, & Bracke, 2015). During crisis there is a reported increase in unemployment rate which is positively correlated with depression rate (Choudhry, Enrico, & Marcello, 2012; Choudhry, Marelli, & Signorelli, 2010).

The findings of this study highlight the impact of COVID-19 global pandemic on psychological well-being of individuals, particularly HCPs and USs. It is our hope that these findings raise awareness amongst policy makers and mental health providers in order to take the necessary measures to attend to psychological wellbeing of individuals during global pandemic. Further studies to investigate the impact of time on mental health are needed.

## Strengths and limitations

To the best of our knowledge, this is the first and largest (4,126 participants, including 1,163 HCP) study in the Middle East that investigated the prevalence of depression and anxiety during COVID-19 pandemic. The large sample size increased the generalisability of these findings. Additionally, the use of previously validated assessment tools is another strength of the study.

However, there are some limitations. There are limited studies that explored the prevalence of depression and anxiety during COVID-19 pandemic worldwide and in the Middle East specifically, a fact that limited our ability to compare our findings with similar healthcare environment and culture. The sample size of ENT physicians’ subgroup was small due to small population in this category nationwide. The impact of time on mental health was not captured here due to the nature of this study and further studies are necessary. It would be useful to repeat the study after the COVID-19 pandemic reach a peak to determine the effect of time on the results. Although depression and anxiety are closely related, depression is almost related to disparate life events, and needs a longer duration in time than the 2-weeks, that are monitored by the PHQ-9 instrument. However, this remark is not valid for anxiety and GAD-7 is relevant to the subject. The above-mentioned remarks may explain the surprising conclusion that USs are more affected and have higher depression and anxiety rates compared to HCPs who are in the centre of the risk and seriously affected by this pandemic disease. Finally, we used an online survey for data collection and therefore, we may have missed some of the targeted population. However, we tackled this by distributing the survey among three different populations and widely used social media.

## 5. Conclusion

Depression and anxiety are prevalent mental problems during COVID-19 pandemic. These mental problems impact the psychological wellbeing of individuals from the entire community including university students, healthcare professionals, and the general population. Females, university students, divorced individuals, healthcare professionals at front-line, and those who are with underlying chronic conditions are at a higher risk. Policymakers and mental care health providers are advised to attend to and provide mental support as needed to vulnerable groups during this pandemic.

## Data Availability

The data that support the findings of this study are available from the corresponding author upon reasonable request.

## Acknowledgments

This study was supported by Isra University (Amman, Jordan).

## Author contributions

AN conceived the study, wrote the methods, conducted the formal analysis, and coordinated the study. AN, EZD, HA, MB, AA, RA, and HMA drafted the manuscript with input from all authors. All authors have been involved in drafting the manuscript or revising it critically for important intellectual content. All authors read and approved the final manuscript.

